# Making Biological Ageing Clocks Personal

**DOI:** 10.1101/2024.02.28.24303427

**Authors:** Murih Pusparum, Olivier Thas, Stephan Beck, Gökhan Ertaylan, Simone Ecker

**Author notes:** Equal Contributions (Gökhan Ertaylan, Simone Ecker).

## Abstract

**Background:** Age is the most important risk factor for the majority of human diseases. Addressing the impact of age-related diseases has become a priority in healthcare practice, leading to the exploration of innovative approaches, including the development of predictors to estimate biological age (so-called “ageing clocks”). These predictors offer promising insights into the ageing process and age-related diseases. This study aims to showcase the significance of ageing clocks within a unique, deeply phenotyped longitudinal cohort. By utilising omics-based approaches alongside gold-standard clinical risk predictors, we elucidate the potential of these novel predictors in revolutionising personalised healthcare and better understanding the ageing process.

**Methods:** We analysed data from the IAM Frontier longitudinal study that collected extensive data from 30 healthy individuals over the timespan of 13 months: DNA methylation data, clinical biochemistry, proteomics and metabolomics measurements as well as data from physical health examinations. For each individual, biological age (BA) and health traits predictions were computed from 29 epigenetic clocks, 4 clinical-biochemistry clocks, 2 proteomics clocks, and 3 metabolomics clocks.

**Findings:** Within the BA prediction framework, comprehensive analyses can discover deviations in biological ageing. Our study shows that the within-person BA predictions at different time points are more similar to each other than the between-person predictions at the same time point, indicating that the ageing process is different between individuals but relatively stable within individuals. Individual-based analyses show interesting findings for three study participants, including observed hematological problems, that further supported and complemented by the current gold standard clinical laboratory profiles.

**Interpretation:** Our analyses indicate that BA predictions can serve as instruments for explaining many biological phenomena and should be considered crucial biomarkers that can complement routine medical tests. With omics becoming routinely measured in regular clinical settings, omics-based BA predictions can be added to the lab results to give a supplementary outlook assisting decision-making in doctors’ assessments.

**Funding:** -

## Introduction

Age is the most important shared risk factor for the majority of human diseases. Hence, there are strong efforts towards attenuating ageing-related disease risk via lifestyle, pharmacological, and clinical interventions to slow or reverse ageing. “Chronological age” (CA), defined by the time passed since an individual’s birth, falls short of reflecting interindividual and environmental differences acting upon a biological system during this period. A critical prerequisite in the endeavour to improve healthy ageing is to quantify an individual’s “wellness,” which covers not only the absence of sickness but also their resilience to future disease, general satisfaction with their health, and having sufficient energy levels for activities that enrich one’s life. While a variety of signals related to individual health and well-being can be collected, validating their contribution to clinically relevant outcomes remains an open issue. The hallmarks of ageing include genomic instability, telomere attrition, epigenetic alterations, loss of proteostasis, disabled macroautophagy, deregulated nutrient-sensing, mitochondrial dysfunction, cellular senescence, stem cell exhaustion, altered intercellular communication, chronic inflammation and dysbiosis^1^. Each hallmark contributes to the ageing process. The major challenge is to dissect the interconnectedness between these hallmarks and their relative contributions to ageing^1^.

To achieve significant progress in the wellness and longevity area to improve human health span, we need to develop tools and methodologies for standard collection, harmonisation, analysis, integration, and interpretation of this information at the individual level. A variety of biological age predictors have been proposed. They share the common approach of using large cross-sectional “healthy” discovery population cohorts with age as a phenotype to construct a predictive model for an organism’s age and subsequently validate it in a separate cohort. Based on the type of molecular data employed, these predictors can be classified into six categories: Telomere length, epigenetic clocks, transcriptomic predictors, proteomic predictors, metabolomics-based predictors, and composite biomarker predictors. Among these, epigenetic clocks are considered to be the most accurate in predicting biological age and other health phenotypes^2^.

Telomeres are repetitive DNA sequences capping chromosomes that shorten every time a cell divides; thus, telomere length (TL) is a conventional marker of biological ageing across various biological domains.^3^ TL has been associated with biological age, wellness, and mortality risk of an individual^4–6^. Furthermore, TL has been proposed for some specific types of cancer^4^ and cardiovascular mortality predictions.^5,6^ However, TL is hard to measure in clinical practice, and recent reports comparing TL with epigenetic clocks on non-symptomatic (or healthy) individuals found TL to be less informative^2,7–9^.

Epigenetic clocks are molecular tools based on 5mC methylation changes to a person’s DNA over time. These modifications, which can be influenced by various factors, including environmental exposures and lifestyle choices, can change over time and, therefore, be used to predict a person’s age.

The “epigenetic clock” premise is to link developmental and maintenance processes to biological ageing, giving rise to a unified theory of the life course of an organism^10^. Epigenetic clocks are used in ageing research to identify potential interventions that could delay or reverse age-related changes and to understand the biological processes underlying ageing. They are also employed to study the relationship between epigenetic changes and various age-related diseases and conditions, such as cancer and cardiovascular disease. Epigenetic clocks are not yet widely used in clinical practice but show promise as a way to measure biological ageing and identify interventions that may be able to improve health outcomes. There is a rapidly growing number of epigenetic clock estimators built for distinct purposes^11,12^, and their application potential, together with the other omics clocks, has also been discussed^13,14^. Proteomics, metabolomics, and multiple clinical biomarker readouts are used to predict biological age, specific disease risks or phenotypes^2^.

Many of those predictors of age and fitness have already been proposed to be used in healthcare practice, and several companies have started to offer direct-to-consumer products in this context^15^. Their product implementation in clinical practice requires rigorous validation and simple and clear recommendations for both the healthcare practitioner and the individual. Hence, despite the large consensus of their premise, there are significant challenges to overcome to transfer scientific health and wellness tools into clinical practice^16^.

In this study, we aim to demonstrate the value of epigenetic clocks, proteomics, metabolomics, and multi-biomarker predictors in a unique longitudinal pilot cohort^17^ where all required data types (clinical parameters, epigenomics, proteomics, metabolomics, along with deep phenotyping and metadata) are available across multiple time points in a 13-month period.

## Methods

### Study design and participants

The IAM Frontier study is a unique longitudinal cohort study that ran for 13 months in 30 healthy individuals, consisting of 15 male and 15 female participants. The 13 months study duration was chosen to cover the seasonal fluctuations that might occur over a one-year period. The study specifically targeted the employees of the research organisation VITO within the age range of 45-59. A major reason for the selection of this group of employees was that as they are part of a research organisation, they are expected to be more open to research-grade technologies and interventions. The age range was selected because the highest prevalence of onset of chronic diseases occurs from the age of 45-65^18^. Individuals were selected based on the following inclusion criteria: not suffering from a chronic disease, diagnosed and currently followed-up by a medical specialist: asthma, chronic bronchitis, chronic obstructive pulmonary disease, emphysema, myocardial infarction, coronary heart disease (angina pectoris), other serious heart diseases, stroke (cerebral haemorrhage, cerebral thrombosis), diabetes, cancer (malignant tumour, also including leukaemia and lymphoma). At monthly visits, a range of samples (whole blood, plasma, urine, stool) were collected and sent to accredited laboratories and comprehensive multi-omics and clinical biochemistry data were assessed. Self-administered questionnaires on, for example, health conditions and physical activity were also completed by the participants. In this article, we analysed data from the IAM Frontier study, which included DNA methylation data, clinical biochemistry, proteomics, metabolomics, and data from physical health examinations.

### Sample collection

The sampling of the IAM Frontier study took place between March 2019 and March 2020 and included the collection of human biospecimen and digital data. During the 13 months of the study, the participants donated blood, urine, and stool samples at monthly visits. These samples were collected after overnight fasting for at least eight hours. The urine and blood samples (in EDTA-, citrate-, and serum-vacutainers) were transported to the clinical laboratory at room temperature within 6 hours after collection. Peripheral blood mononuclear cells (PBMC) were isolated from EDTA-blood samples, and PBMC pellets were stored at −80°C till the DNA extraction. At monthly visits, clinical tests and health examinations such as blood pressure, body height, weight, and abdominal circumference measurements were performed by accredited labs and appointed doctors. At bi-monthly visits, plasma samples were taken, and omics (proteomics and metabolomics) measurements were assessed. At months one, six, and 13, the PBMC samples were used to measure DNA methylation. At month 13, only 20 participants were able to donate samples due to the start of the COVID-19 pandemic. Table 1 presents an overview of the sample collection. All samples have been collected in accordance with the applicable Belgian regulations regarding the use of human body material for scientific research (Belgian Law on use of human body material, 2008) and the Belgian Royal Decree on biobanks (*Het Koninklijk Besluit betreffende de biobanken. Belgisch Staatsblad 05.02.2018. Brussels (2018)*).

**Table 1:**
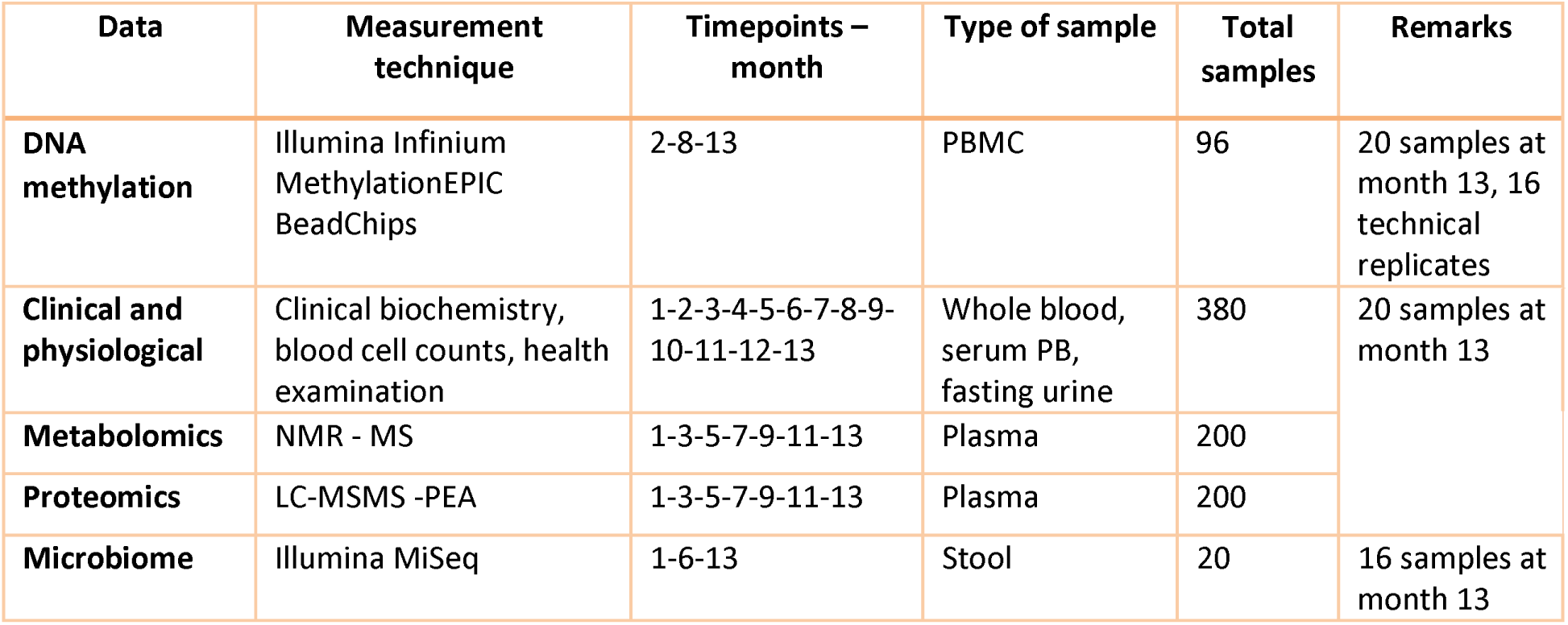
Data collection of IAM Frontier study.

### DNA methylation assay

The DNA methylation assay was carried out using Diagenode Epigenomic Services (Vienna, Austria, Cat No. G02090000). PBMC samples were sent for DNA methylation profiling using the Illumina Infinium MethylationEPIC array BeadChip (850K) platform to analyse the methylation status of more than 850,000 CpGs per sample. This microarray covers 196% of CpG Islands and 99% of annotated RefSeq genes. We performed the DNA methylation data pre-processing and the corresponding details can be found in Supplementary Document Section 1.

### Analysis of epigenetic clocks and predictors

We applied 29 different epigenetic age and health trait predictors (see Figure 2). The Skin & Blood Clock^19^, Multi-tissue Clock^20^, HannumAge^21^, DNAmTL^22^, PhenoAge^23^, GrimAge^24^, GrimAge DNAmPACKYRS and GrimAge protein levels (DNAmADM, B2M, CystatinC, GDF15, Leptin, PAI1, TIMP1) were obtained by Steve Horvath’s DNA Methylation Age Calculator available on http://dnamage.genetics.ucla.edu/. The MethylDetectR predictions (Age, Alcohol, BMI, HDL, BodyFat, Waist:Hip Ratio and Smoking)^25^ were calculated using the code available at https://zenodo.org/record/4646300. The methylation Pace of Age (mPoA)^26^ was estimated using the code available at https://github.com/danbelsky/DunedinPoAm38. The MetaClock^27^ code was received by e-mail from the author Morgan E Levine. EpiTOC scores were calculated using the code available in the corresponding publication.^28^ EpiTOC2^29^ scores were calculated using the code available from https://doi.org/10.5281/zenodo.2632938, and MiAge^30^ scores were calculated using the code available from http://www.columbia.edu/~sw2206/softwares.htm. Alcohol predictions^31^ were generated using the dnamlci R package available from https://github.com/yousefi138/dnamalci. Elliot’s smoking score^32^, Zhang’s smoking score^33^ and EpiSmokEr’s smoking status^34^ were obtained using the R package EpiSmokEr available at https://github.com/sailalithabollepalli/EpiSmokEr.

**Figure 1:**
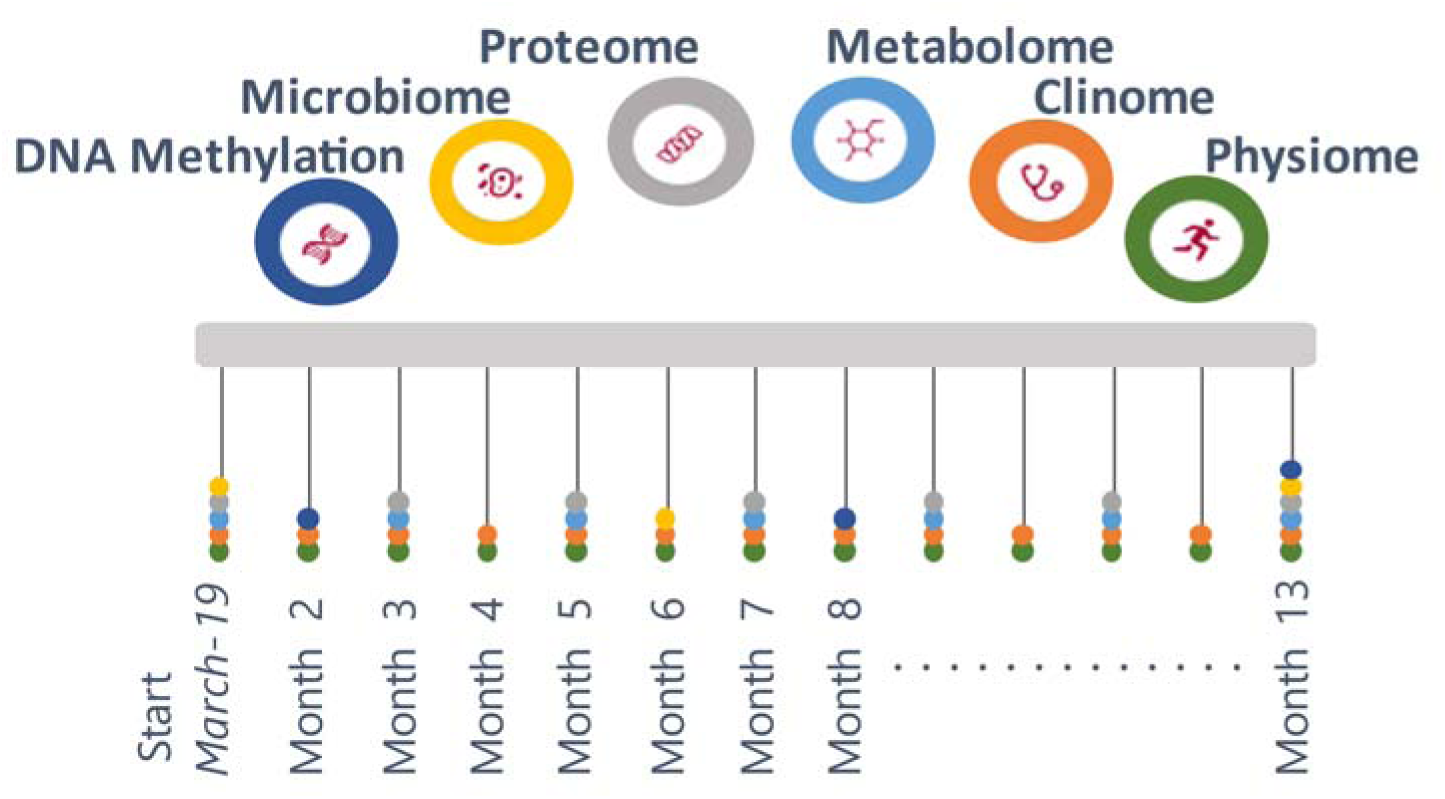
Study design of the IAM Frontier. Longitudinal comprehensive data were collected over the course of the study including monthly physiological and clinical biochemistry data (13 time points), bimonthly proteomics and metabolomics data (7 time points), and six-monthly DNA methylation and microbiome data (3 time points). The study participants consist of 15 males and 15 females, with a (chronological) age range of 45 – 59 years old.

**Figure 2:**
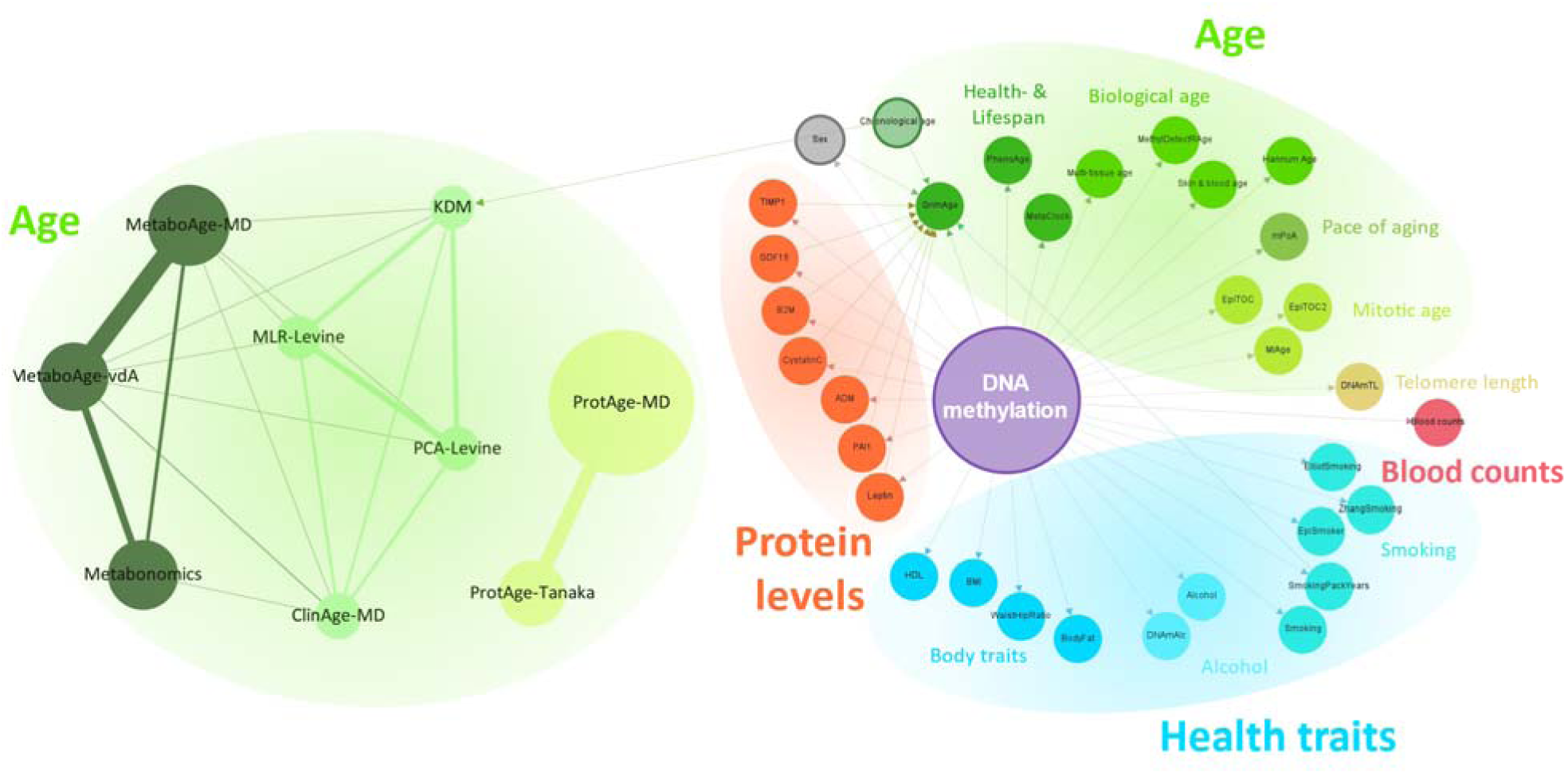
The overview of published clinical and omics ageing clocks and predictors discussed in this article. The predictors are grouped based on their purpose on predicting age, blood counts, health traits, and protein levels. Each node represents one predictor connected to another. In the left-hand panel, the edges represent the ratio of shared parameters, e.g., there are 26 shared parameters between the MetaboAge-vdA clock and the MetaboAge-MD clock, hence the thickest edge. The size of each node represents the total number of parameters used in each clock. The right-hand panel shows all DNA methylation predictors used in this study. Edges indicate the parameters used for each predictor. Due to the high numbers of DNA methylation sites used by the predictors, nodes and edges are shown in standard sizes, with no relation to the number of features used or shared.

GrimAge, HannumAge, MethylDetectRAge, mPoA, as well as the smoking, alcohol and health trait predictors, were originally developed for whole blood measurements but have been shown to work well with PBMCs in our study and others^35–38^. Table 2 shows the list of clocks implemented in this study including clocks predicted from clinical, metabolomics, and proteomics data (see the next section).

**Table 2:** List of applied clinical and omics ageing clocks. Rows with colours refers to clocks that are further investigated: yellow, red, and blue rows correspond to the clocks that were estimated using clinical, proteomics, and DNA methylation data, respectively.

| Clock | Reference | Remarks |
| --- | --- | --- |
| **MLR-Levine**<sup>42</sup> | Levine, ME. 2013. Gerontol A Biol Sci Med Sci. | Predicts chronological/biological age |
| **PCA-Levine**<sup>42</sup> |  | Predicts chronological/biological age |
| **KDM** | Klemera, P. and Doubal, S. 2006. Mech Ageing Dev.<br>Levine, ME. 2013. Gerontol A Biol Sci Med Sci. | Predicts chronological/biological age |
| **ProtAge-Tanaka**<sup>43</sup> | Tanaka, T., et al. 2018. Aging Cell. | Predicts chronological/biological age |
| **ProtAge-MD**<sup>44</sup> | Macdonald-Dunlop, E., et al. 2022. Aging. | Predicts chronological/biological age |
| **ClinAge-MD**<sup>44</sup> | Macdonald-Dunlop, E., et al. 2022. Aging. | Overestimated BA (median BA 750 years), not shown |
| **MetaboAge-MD**<sup>44</sup> |  | Overestimated BA (median BA 600 years), not shown |
| **MetaboAge-vdA**<sup>45</sup> | van den Akker, EB., et al. 2020. Circ Genom Precis Med. | Underestimated BA (median BA –5000 years), not shown |
| **Metabonomics**<sup>46</sup> | Hertel, J., et.al. 2016. Journal of Proteome Research. | Missing variables (only 22% vars. are available), not shown |
| **Skin & blood clock**<sup>19</sup> | Horvath, S., et al. 2018. Aging. | Predicts chronological age |
| **MethylDetectR Age**<sup>25</sup> | Hillary, R. and Marioni, R. | Predicts chronological age |
| **Multi-tissue clock**<sup>20</sup> | Horvath, S. 2013. Genome Bioloy. | Predicts chronological/biological age |
| **Hannum clock**<sup>21</sup> | Hannum, G., et al. 2013. Molecular Cell. | Predicts chronological/biological age |
| **mPoA**<sup>26</sup> | Belsky, D., et al. 2020. eLife. | Predicts the pace of ageing |
| **PhenoAge**<sup>23</sup> | Levine, M., et al. 2018. Aging. | Predicts health- and lifespan |
| **GrimAge**<sup>24</sup> | Lu, A., et al. 2019. Aging. | Predicts health- and lifespan |
| **Metaclock**<sup>27</sup> | Liu, Z., et al. 2020. Aging Cell. | Predicts mortality |
| **EpiTOC**<sup>28</sup> | Yang, Z., et al. 2016. Genome Biology. | Predicts mitotic age |
| **EpiTOC2**<sup>29</sup> | Teschendorff, A.E. 2020. Genome Medicine | Predicts mitotic age |
| **MiAge**<sup>30</sup> | Youn, A., and Wang, S. 2018. Epigenetics. | Predicts mitotic age |
| **DNAmTL**<sup>22</sup> | Lu, A., et al. 2019. Aging. | Predicts telomere length |

We also predicted blood counts using minfi^39^ and the Reinius reference dataset^40^ as well as IDOL using the Salas reference dataset^41^ with the code available from https://github.com/immunomethylomics/FlowSorted.Blood.EPIC.

The correlation between predicted and chronological age was calculated using Pearson’s correlation. We excluded technical replicates from the calculation of correlation coefficients.

For the longitudinal analyses of epigenetic predictions, we used the 16 technical replicates available from our study to estimate technical variation. Two technical replicates were generated within each of the three DNA methylation time points, and ten additional technical replicates of the two previous time points were generated at the third time point, providing detailed insight into technical variation influencing the predictions. We used the maximum absolute difference observed between technical replicates as a threshold to define potential biological differences across time points after also adding the chronological time that passed between time points. For example, the maximum absolute differences observed for the Skin & Blood Clock across technical replicates was 2.56 years (the reported median error in blood is 2.5 years^19^), and the threshold we used represents the maximum differences observed among technical replicates plus the chronological time that passed between the time points which are maximum 343 days (0.94 years).

For the global analyses of our dataset and the identification of biological outliers of interest based on our cohort, we introduced a Mean Absolute Deviation (MAD) threshold of +/-2*MAD and +/-3*MAD across all biological replicates.

### Analysis of Clinical, Metabolomics, and Proteomics Clocks

The monthly clinical and bi-monthly metabolomics as well as proteomics data were used for predicting longitudinal biological age using several published calculators. We applied the multiple linear regression (MLR) model and principal component analysis (PCA) developed by Levine *et al*.^42^ –they will be referred to as MLR-Levine and PCA-Levine– to the clinical data that consist of samples from 12-13 time points per individual. The model prediction involves both clinical and physiological measurements, including total cholesterol level, glycated haemoglobin, C-reactive protein, systolic blood pressure, forced expiratory volume (FEV), and cytomegalovirus (CMV). In the IAM Frontier data, FEV and CMV were not measured, but we imputed the values with the corresponding median from the original study^47^. The same clinical variables as in the PCA-Levine model were further used to predict the biological age using the Klemera and Doubal method (KDM).^48^ Unlike the MLR-Levine and the PCA-Levine, KDM also incorporates chronological age in their estimation procedure. Similarly, we also computed other biological age predictions based on similar clinical biochemistry variables developed by McDonald-Dunlop et al.^44^

The proteomics and NMR metabolomics data consist of samples from six to seven time points per individual, where the samples were sent to the laboratory in four different batches. For the proteomics data, batch correction normalisation was done prior to the analysis to reduce the technical variation between batches/plates^49^. We performed PCA and multidimensional scaling (MDS) analyses for the metabolomics data, where we did not observe any batch effects, so the raw metabolite abundances were used. In both datasets, there are twenty subjects with technical replicates spread across different time points. We performed a procedure for selecting the samples (between the originals and the replicates) by computing cosine similarity coefficients for all samples. The samples with the closest similarity to the rest of the individuals’ measurements were selected. Further, we predicted the biological age using other published proteomics and metabolomics clocks: ProtAge-MD (from McDonald-Dunlop et al.^16^), MetaboAge-MD, ProtAge-Tanaka, MetaboAge-vdA, and Metabonomics^43–46^.

### Software tools and programming language

The network (Figure 2) was created in Gephi version 0.9.5^50^ and Cytoscape version 3.8.2^51^. All data analyses were conducted in the R statistical environment version 3.5.1, 4.1.0 and 4.1.3.

## Results

From March 2019 to March 2020, the IAM Frontier study collected monthly samples and data (via online questionnaires and wearable sensors) from 30 healthy (no diagnosed chronic diseases, no self-reported illnesses except hypertension) and highly motivated individuals. An intake interview was performed to check whether the volunteers fit the inclusion and exclusion criteria (health, age, sex balance) and to assess their motivation to join and stay within the study, as the study required monthly site visits for sample collection, continuous wearing of sensors and weekly questionnaires. All individuals were followed up with study doctor visits to inform them about their health status (from the clinical grade biomarkers) as well as provided with genetic counselling with certified personnel when necessary. The study design is illustrated in Figure 1.

In this study, we first explore the global pattern of the values resulting from a multitude of ageing clocks and examine their utility in predicting personal wellness, health, and biological age (BA). These clocks were computed for all the IAM Frontier individuals, utilising their clinical biochemistry and physiological data (such as blood pressure, weight, and height), DNA methylation, metabolomics, and proteomics measurements. Further, we investigate how these clocks predict outcomes across different time points, leveraging the longitudinal nature of the IAM Frontier study. Individuals with interesting findings are subjected to further investigation to demonstrate the importance and the relevance of the omics clocks as personal health assistants, capable of monitoring and assisting the clinical examination and diagnosis.

Figure 2 shows the overview of ageing clocks that can be estimated from various omics measurements. We categorise these clocks according to their utility in predicting 1) BA, 2) blood counts, 3) health traits such as smoking status, alcohol consumption and body mass index (BMI), and 4) plasma protein levels. By using DNA methylation data, 13 clocks aim to predict age, one clock to predict blood counts, 11 clocks to predict health traits, and seven clocks to predict plasma protein levels. In addition, three clinical biochemistry clocks, three metabolomics, and two proteomics clocks can also be used for predicting BA from clinical biochemistry data, NMR metabolomics, and Olink proteomics measurements. In this study, we focus on investigating the ageing clocks that are particularly useful in predicting BA.

In general, in each data type, different ageing clocks use a different set of features. The features used in ageing clocks will further be referred to as variables. Figure 2 shows that there are some variables that are shared and utilised in more than one clock, as demonstrated by the edges between clocks (see MLR-Levine and KDM). It is also possible that these variables are shared across different data types, for example between the MLR-Levine and the Metabo-MD where albumin is utilised in both clocks. Albumin and several other variables are present in the clinical biochemistry and metabolomics data; they refer to the same clinical compounds but are measured by different technologies. Since our study focuses on ageing clocks that aim to predict BA, it is also worth noting that there are several omics ageing clocks that include chronological age (CA) in their algorithm, for example, GrimAge and KDM. GrimAge also includes sex and plasma protein levels predicted from the DNA methylation data in its calculation.

Epigenomic, metabolomic and proteomic data are high dimensional. The IAM Frontier study collected the DNA methylation measurements of more than 850,000 sites per sample using Illumina’s Infinium MethylationEPIC BeadChips as well as 1,068 proteins and 249 metabolites were measured in the metabolomics and proteomics data of IAM Frontier, respectively. Large numbers of variables are used in ageing clocks working with these data types. Although several epigenetic age clocks based on only a few loci have been proposed^26,52^, they tend to be less accurate. The most widely used epigenetic clocks and predictors incorporate between 71^21^ and 1030 CpGs^24^. In comparison, ageing clocks based on clinical biochemistry data or other omics data (metabolomics, proteomics) require nine to 203 parameters^42,44^.

The clocks we implemented are presented in Table 2. Some of the clocks were not further investigated due to showing unrealistic predictions, unavailable model coefficients (e.g., the model intercept coefficients are not published in the MetaboAge-MD clock), or unavailable variables (e.g., due to differences in the omics technology, around 80% of the required metabolites used in the Metabonomics clock were not measured in the IAM Frontier dataset)^22,26–30,44–46^. Different sets of variables with different sizes are involved in each clock. The included algorithms do not constitute an exhaustive list but were selected in their applicability to the IAM Frontier study with high confidence due to data availability and matching underlying assumptions.

Although the study’s main objective is focused on BA prediction, CA is still incorporated in some analyses. While CA may not provide the most robust description of human ageing, it offers the most convenient way to calculate age and is perceived as a standard measure of ageing. Stratification by CA group is also often done, for example in clinical reference intervals^53,54^. For this reason, we believe it is valid to compare the predictions of BA with the subjects’ CA; see the Pearson correlation coefficients in Table 3. All the clocks shown in Table 3 are significantly correlated with CA; MethylDetectR and the Skin & Blood clock give the highest correlation coefficients, *R*=0.90 and *R*=0.87, respectively. This is not unexpected, as both clocks were developed to predict CA. The kernel densities of the predicted BA as well as CA are shown in Figure 3. The predicted Skin & Blood clock age gives a similar kernel density as the CA, both in the position as well as the shape. From this perspective, we may suggest this clock as the best tool to measure individuals’ chronological ageing. However, the correlation between chronological age and predicted age among all measured biological clocks is the highest for MethylDetectRAge, another chronological age predictor (Pearson’s *R*=0.91), although its predictions appear to be slightly shifted towards older ages in our dataset. GrimAge, with *R*=0.85, gives a position of the kernel similar to the chronological age, but with a different shape. GrimAge aims to predict lifespan and healthspan, and as a remark, it also incorporates chronological age in its calculation, unlike the other clocks which are solely based on DNA methylation markers, other omics, or clinical measurements. The same remark also applies to KDM-Levine clock, with *R*=0.76 and an identical position of the kernel density as compared to the chronological age.

**Figure 3:**
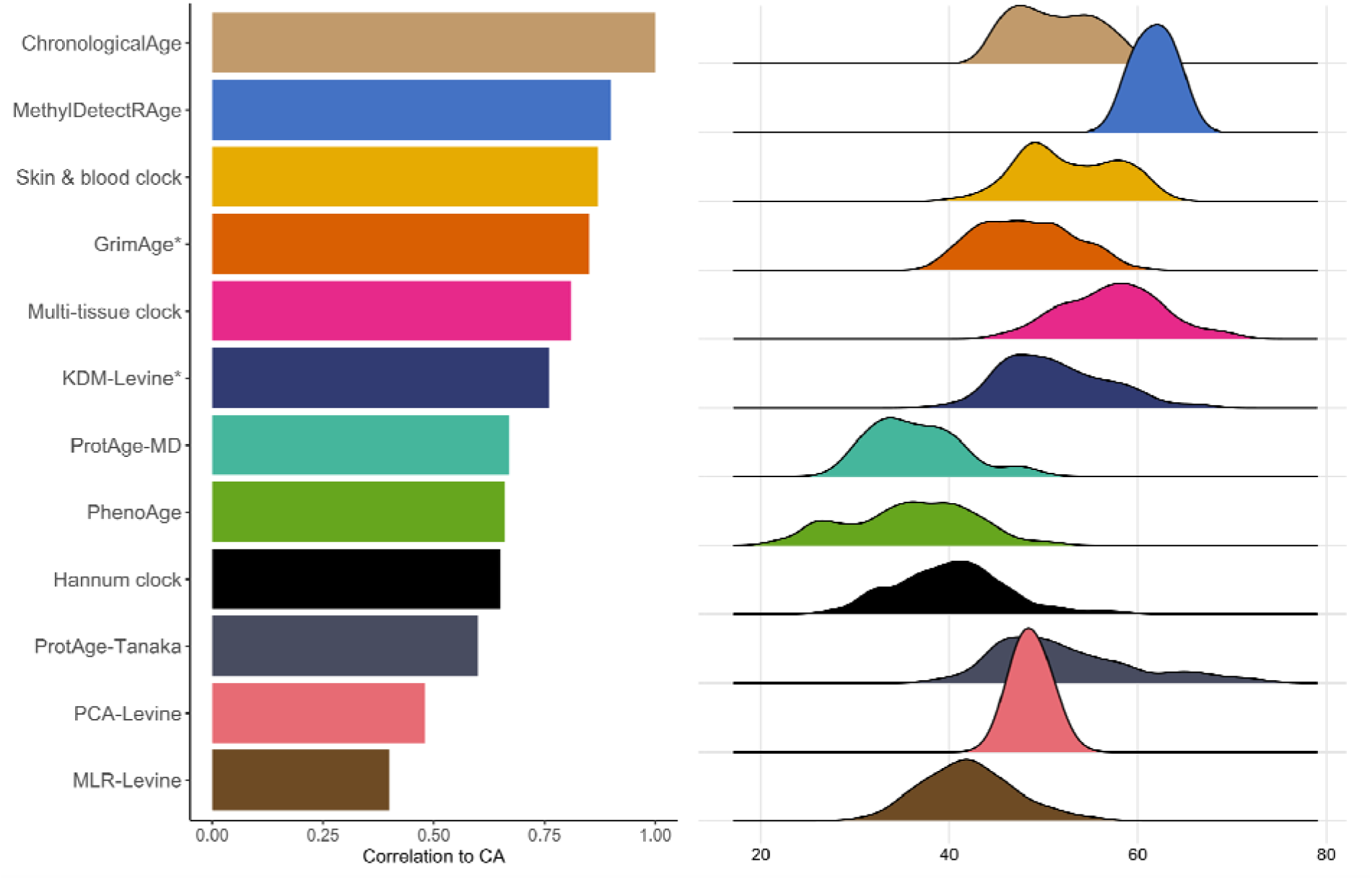
Kernel density plots of BA predictions from several ageing clocks and their correlations with the chronological age (CA).

**Table 3:** Correlation between predicted age and chronological age. Significant correlations are displayed in bold (at p<0.05). *GrimAge and KDM incorporate chronological age in their calculation.

| Predictor | Corr. coefficient | p-value |
| --- | --- | --- |
| **MethylDetectRAge** | **0.90** | **2.0E-30** |
| **Skin & blood clock** | **0.87** | **3.5E-26** |
| **GrimAge** | **0.85** | **1.1E-23** |
| **Multi-tissue clock** | **0.81** | **1.6E-19** |
| **KDM** | **0.76** | **2.2E-16** |
| **ProtAge-MD** | **0.67** | **2.2E-16** |
| **PhenoAge** | **0.66** | **3.8E-11** |
| **Hannum clock** | **0.65** | **4.8E-11** |
| **ProtAge-Tanaka** | **0.60** | **2.2E-16** |
| **MetaClock** | **0.56** | **8.7E-08** |
| **PCA-Levine** | **0.48** | **2.2E-16** |
| **MLR-Levine** | **0.40** | **1.4E-15** |
| **ClinAge-MD** | **0.37** | **1.2E-13** |
| **EpiTOC** | **0.25** | **2.4E-02** |
| MiAge | 0.20 | 7.6E-02 |
| **MetaboAge-vdA** | **0.18** | **8.6E-03** |
| mPoA | 0.18 | 1.1E-01 |
| MetaboAge-MD | 0.08 | 2.6E-01 |
| EpiTOC2 | 0.04 | 7.4E-01 |

We performed different data exploration approaches to compare the BA predictions of each individual who participated in the IAM Frontier study. At each BA predictor, we observed smaller within-person predictions at different time points (focusing on *inter-individual variability*) than the between-person predictions at the same time point (focusing on *intra-individual variability*). The small inter-individual variabilities essentially show that the predictions within individuals are closer to each other than between the peers; this observation is similar to the characteristics of common clinical biomarkers when they are measured longitudinally. Therefore, it is reasonable to analyse the results by looking at the differences between individuals, and further explore the fluctuation of the predictions within each individual separately. In Figure 4A, for example, age accelerations which correspond to the difference between predicted BA and CA are shown at time points two, eight, and 13; the time points that are shared between all omics and clinical data. Fairly distinct predictions are observed for each clock, and fluctuations are also seen between time points. Within the same clock, we observe that the between-individual fluctuations are larger than the within-individuals’. The age acceleration predictions of GrimAge, the Skin & Blood clock, KDM-Levine, PCA-Levine, and ProtAge-Tanaka are close to zero, i.e. their predicted BA values are close to the corresponding CAs. Referring to the estimated kernel density in Figure 3, the predicted values of these clocks are similar to the CA with reference to the location. The predictions observed in the clinical and proteomics data for all time points are shown in Figure 4B.

**Figure 4:**
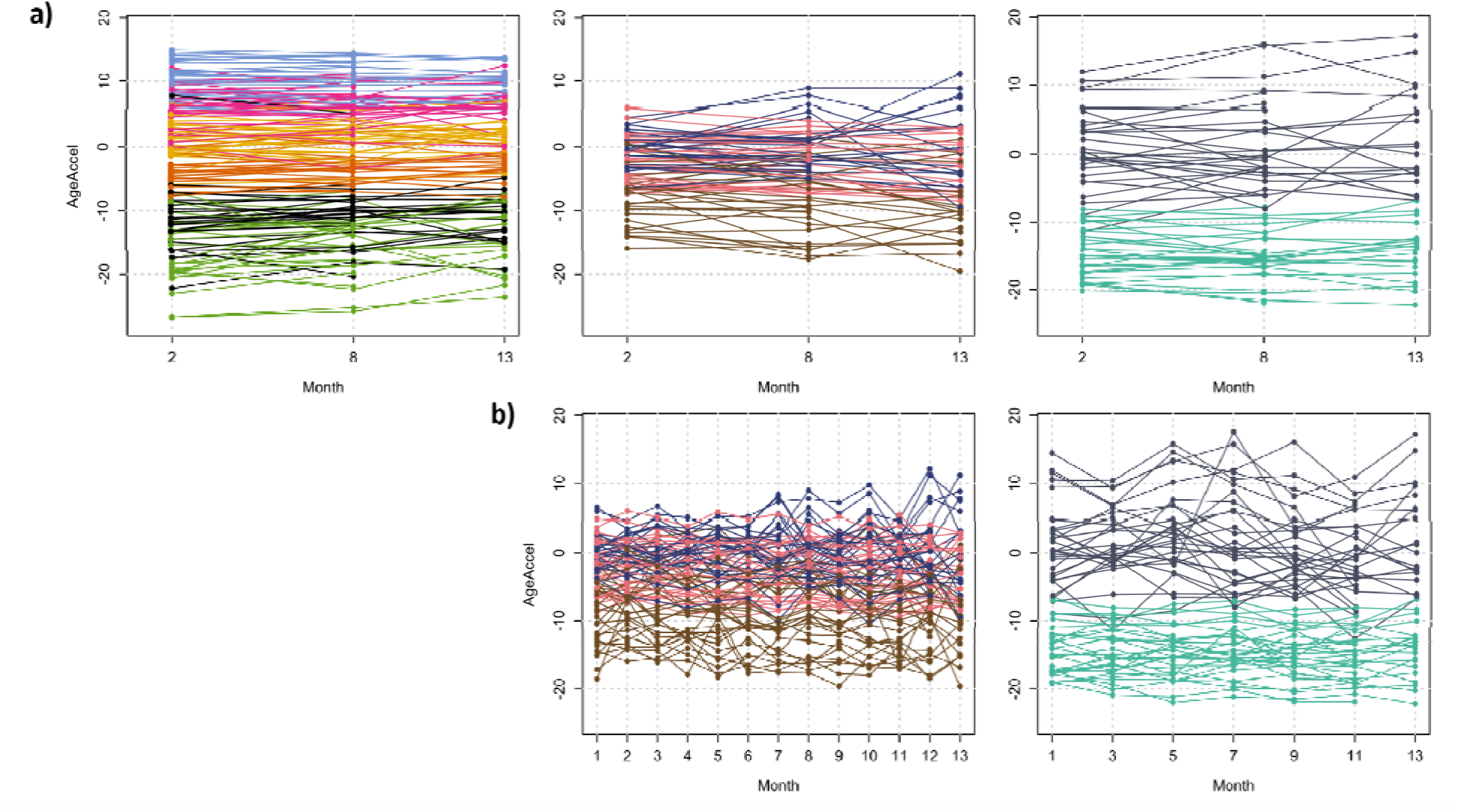
Age acceleration of all IAM Frontier participants in different clocks. (A) Age acceleration based on the Multi-tissue clock (pink), Skin & Blood clock (yellow), PhenoAge (green), GrimAge (orange), MethylDetectR (light blue), and Hannum clock (black). (B) Age acceleration based on MLR-Levine clock (brown), PCA-Levine clock (light pink), and KDM clock (blue). (C) Age acceleration based on ProtAge-Tanaka (grey) and ProtAge-MD (tosca).

We continue the analyses by investigating the BA and health traits predictions in all IAM Frontier participants. Due to the small sample nature of the IAM Frontier study, we are able to examine the individual predictions resulting from all clocks. Figure 5A shows the ordered heatmaps of the predicted BAs as well as the health trait predictions in all individuals at time points two, eight, and 13. These were selected because the results of all clocks are available at these time points, see Table 1. In Figure 5B, the MAD thresholds were computed and were used to give the colour annotations. Therefore, only deviating predictions appear coloured in the figure. We can observe distinct patterns for ID06, ID08 and ID27, detailed below. In Figure 6B, we show the clinical laboratory profiles of these individuals.

**Figure 5:**
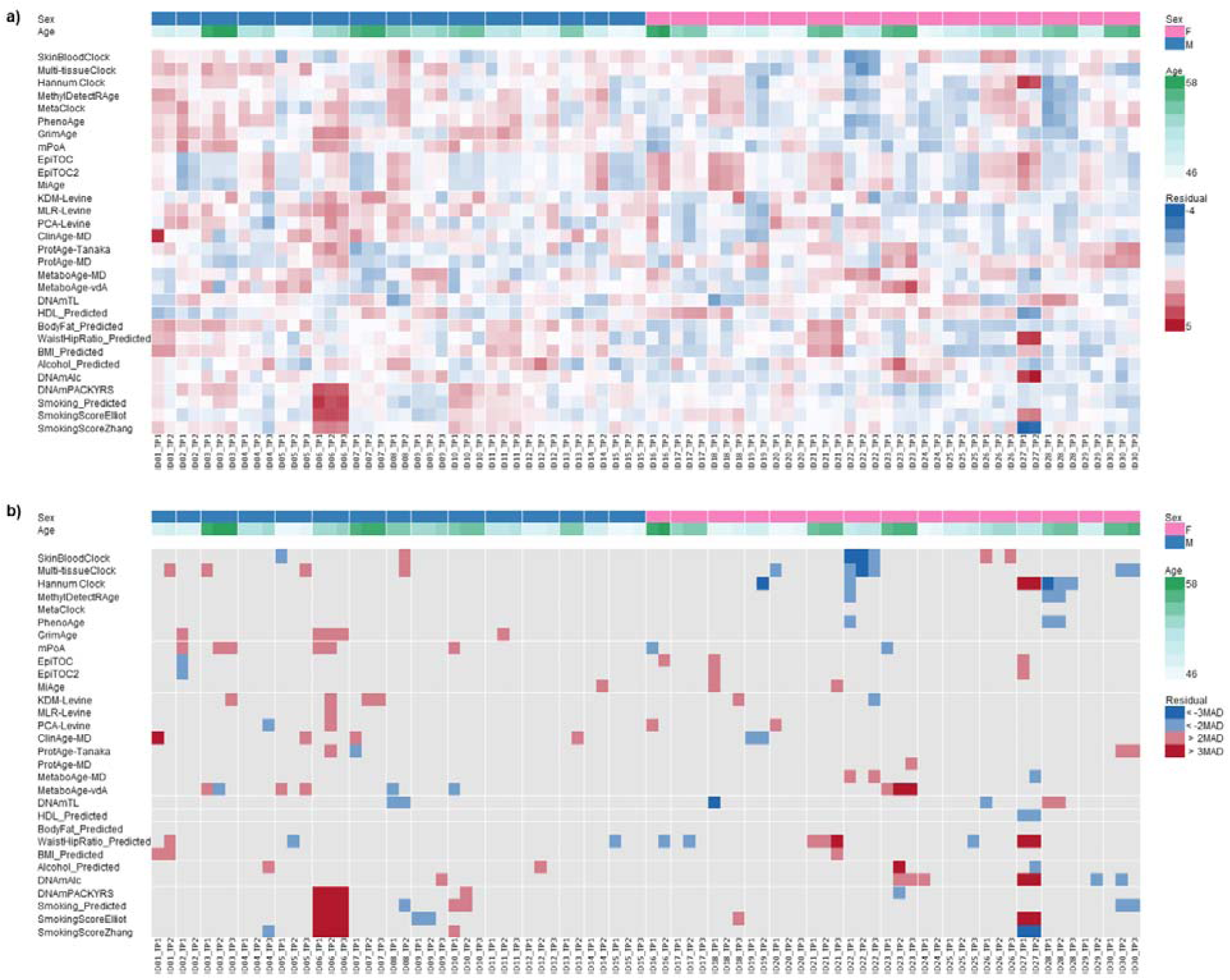
a) Ordered heatmap. ID06 has higher predicted smoking scores and ID27 is an outlier in several predictions. A slight sex effect is also visible. b) Reduced ordered heatmap based on MAD thresholds.

**Figure 6:**
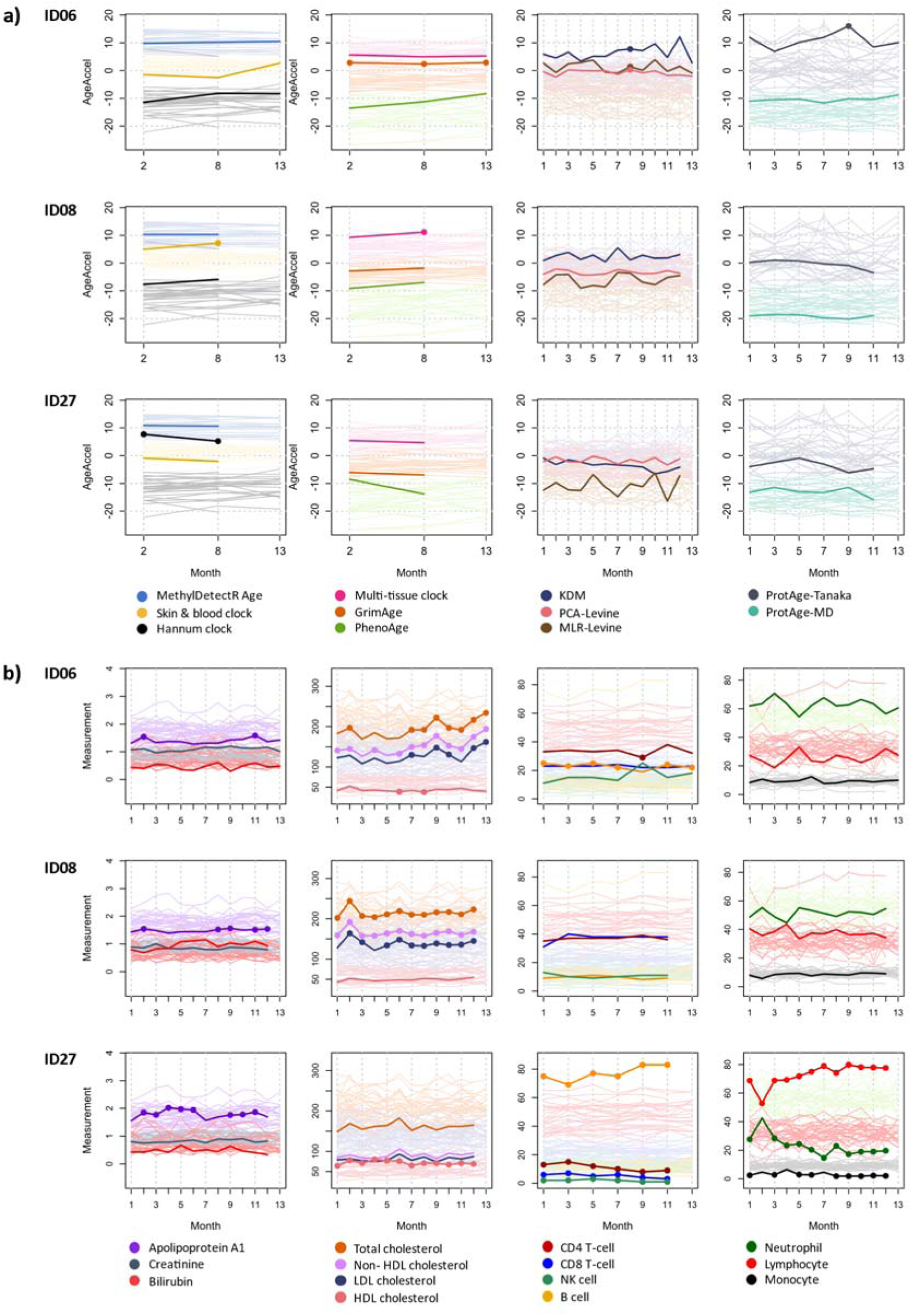
**a)** Age acceleration of ID06, ID08, and ID27. Prediction values that are significantly different from the rest of the cohort are marked with full circles. **b)** Clinical and blood cell profiles of ID06, ID08, and ID27. Measurements that are outside the cohort reference intervals are marked with full circles. The blood cell counts are shown in percentages.

ID06 shows very high smoking predictions. ID06 is a current smoker, indeed, and hence has a higher risk of developing cardiovascular diseases (CVD) and diabetes (Supplementary Figure 3 of CVD and diabetes disease risk scores). From the individual plots in Figure 6A, we also observe that the BA predictions of ID06 increase over time for some of the ageing clocks, such as in the Skin & Blood clock and PhenoAge. This individual was also biologically older at all time points according to the KDM and ProtAge-Tanaka clocks. However, the clinical laboratory profiles of ID06 show an increasing trend of the lipid measurements, in line with this individual’s increased risk of CVD and diabetes. Interestingly, ID06’s B-cell percentages are higher than the normal range, and the CD4 T-cell and lymphocyte counts are low. This could show that this individual might also have problems with antibodies and the immune system. In the questionnaires, ID06 did not report major health burdens except that this individual felt having low energy, tense muscles, and constant fatigue, with the latter two commonly reported among the study participants.

ID08 appears as the only individual with predicted shortened telomeres across both measured time points based on the DNAmTL clock, as seen in Figure 5B. This result may indicate premature ageing. In addition, in almost all the applied epigenetic clocks, this individual was also predicted to be epigenetically older at the second time point compared to the first (see Figure 6A). The lipid (cholesterol) profiles of ID08 (see Figure 6B) show an alarming status; the measurements are out-of-normal range at almost all time points. Other clinical parameters are still within the normal limits, although the percentages of CD8 T-cell and lymphocyte are high compared to the peers. Based on the weekly questionnaires, during the course of this study, this individual experienced constant fatigue, diarrhoea, numbness, indigestion, sleeping problems, stuffy nose, and blurry vision, see Supplementary Figure 5. Although ID08 does not appear to report all of these health burdens at a significantly higher level than the other study participants, together with the rapid epigenetic ageing and high lipid profiles observed, they might provide additional support to the premature ageing condition predicted by the DNAmTL clock.

ID27 appears to be different in our analyses in many ways, although most of the epigenetic, proteomi and clinical ageing clocks predict this individual to be in a healthy state – healthier and biologically younger than the peers. However, ID27 shows very strong outlier BA predictions corresponding to the Hannum clock as well as very different smoking and health trait predictions. It is the only subject with a completely distinct Hannum age acceleration compared to the peers at all time points, epigenetically younger at the second time point as compared to the first across all clocks, and with conflicting smoking predictions (this individual was not a current smoker). The BA predictions of the clinical and proteomics clocks also show a general decreasing trend over time, where ID27 was biologically younger than chronologically, according to some of those clocks. In addition, from the DNA methylation and proteomics analyses, this subject is discovered to be far outside the main cluster in the corresponding PCA plots (Supplementary Figure 4a and 4b). When looking at DNA methylation predicted blood cell composition, we observe a strongly increased proportion of B cell (Supplementary Figure 6). The finding is confirmed by actual blood counts showing the same abnormalities, clearly indicating a hematological problem. This explains why ID27’s age predictions appear as extreme outliers according to Hannum’s clock, as it is sensitive to blood composition changes, and may also explain the conflicting and unexpected outlier predictions obtained for several health traits such as smoking and alcohol intake, among others. Throughout the study period, ID27 reported constant medium backpain, a slight headache and blurry vision.

As the final analysis, we present an unsupervised heatmap and clustering of all predictions and actual measurements of our study in Figure 7. This analysis shows potential patterns arising when all clocks and measurements are included and how they may be related to each other. For example, predictions from epigenetic clocks with high Pearson correlation coefficients with CA (MethylDetectRAge, Skin & blood clock, GrimAge) are indeed clustered together with CA, showing a strong linear relationship and possibly similar predictive mechanism. However, clinical, proteomics, and metabolomics-based clocks stand apart from this cluster, suggesting their predictive mechanisms may be distinct despite their significant correlations with CA.

**Figure 7:**
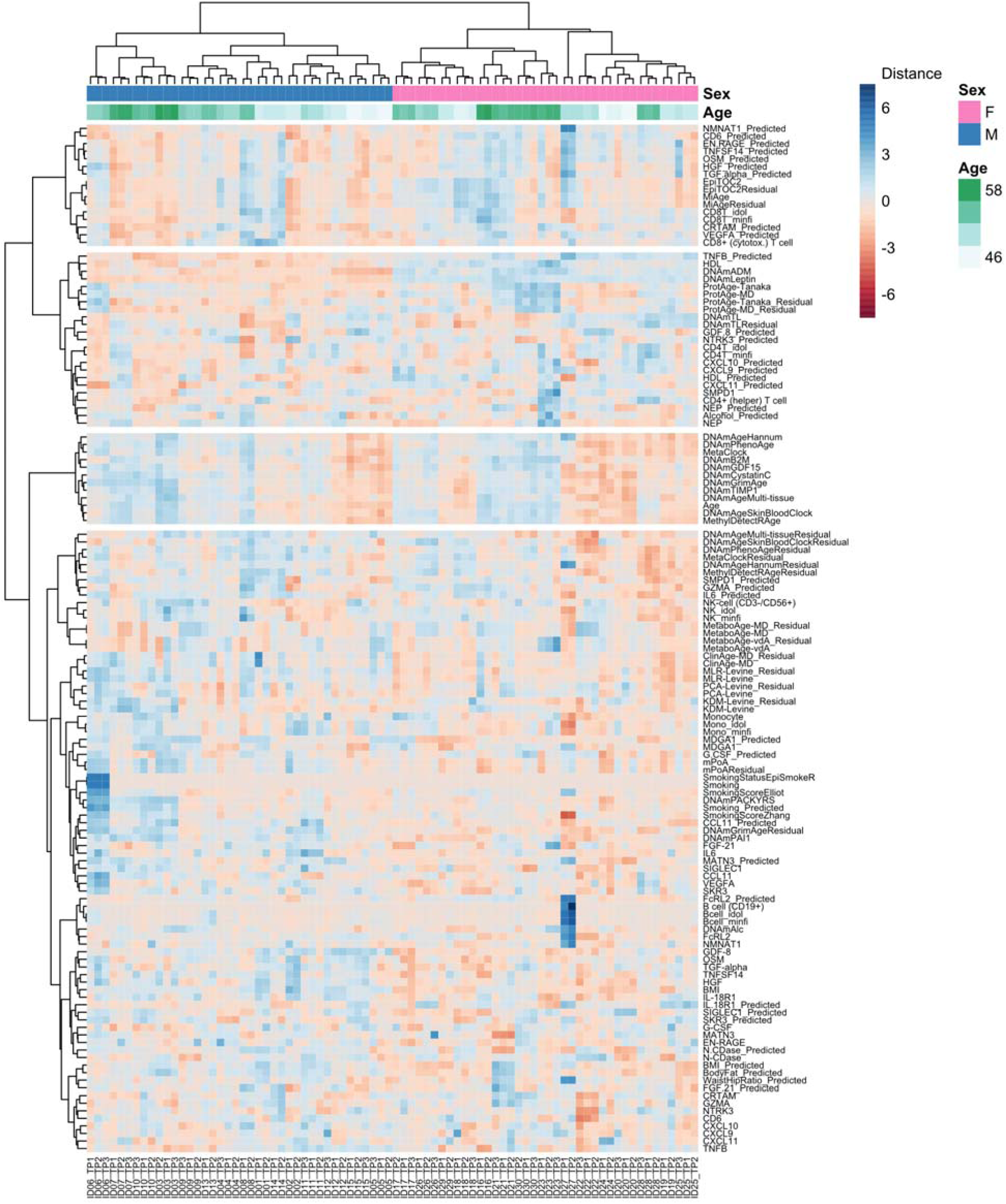
Unsupervised heatmap of the estimates of all clocks and predictors together with the actual measurements. All participants in three DNA methylation time points are included. The clustering based on the Manhattan-Ward D distances is also showed, annotated by participants’ age and sex.

Also in Figure 7, ID06 and ID27 are clearly seen to be different in several indicators, especially in the blood cells prediction. Our results for ID06, ID08, and ID27 as described above illustrate that the deviation of human biological ageing can be discovered when a comprehensive analysis within the biological age prediction framework is done. The IAM Frontier study followed a communication strategy to discuss these findings, or so-called incidental findings (IF) including genetic alterations back to the IAM Frontier participants^55^. Different clocks can show various results, and additional analyses on further assessment types, such as clinical laboratory tests and reports on personal health problems, can complement and enhance the acquired insight.

## Discussion and conclusion

Age stands as the principal risk determinant for ailments, impairments and diseases. The pursuit of mitigating age-related illnesses and extending the healthful years of life has led to the innovative idea of directly targeting the ageing process to restore physiological functionality. Realising this ambitious goal necessitates the precise assessment of biological age and the pace of ageing at the molecular level. Propelled by the latest breakthroughs in high-throughput omics technologies, a novel suite of tools has emerged for the quantitative analysis of biological ageing. By leveraging data from various domains such as epigenomics, proteomics, and metabolomics, and employing machine learning techniques, “biological ageing clocks” have been constructed. These clocks have proven their ability to pinpoint potential biomarkers of biological ageing, offering unprecedented insights into the ageing process.

A recent review paints today’s landscape of biological age (BA) prediction algorithms using omics technologies^56^. These methods were claimed to better represent the biological state of an organism than the chronological age (CA). Ageing clocks for BA are also starting to appear as commercial products, serving as a window into the personal health status. In this manuscript, we use a unique deep-phenotyping dataset that allows us to perform and analyse multiple BA predictors using different data types side by side. The longitudinal nature of the data enables us to study the individuals’ fluctuations between time points, exploring their changes in BA predictions over time. We optimise our analyses using the extensive set of technical replicates (see Table 1) generated to estimate technical variation in our dataset and design our analyses correspondingly (see Methods and Supplementary Material Section 1).

The BA analyses in the IAM Frontier data shows that BA predictions can fluctuate over time; both an increase and a decrease might happen depending on the biological conditions rather than monotonically increase by time, as in CA. Repeated measurements over time are highly valuable, as outliers can also be caused by normal biological fluctuations. In our study, most BA predictions seem to be “stable”, that is, within the expected range of variability across the available time points. However, there are individuals whose predictions do change more than expected over time, and where this is the case, they are often consistent across multiple BA predictors and time points. A comprehensive exploration of the BA prediction differences between individuals of our study leads to three individuals with distinct BA predictions as compared to their peers.

The BA predictions of ID06, ID08 and ID27, give a good example where information from different data types can not only support the results of standard laboratory tests but also provide additional insight. For example, the lipid laboratory test of ID08 shows a worrying result where almost all measurements fall outside the normal ranges. By applying different BA predictors, we discovered that this individual is also predicted to have significantly shortened telomeres across all time points, an indicator of potentially accelerated ageing.

Another important finding is that despite not reporting major or more severe symptoms than the peers, ID27 shows strongly altered epigenetic predictions which are reflected in blood cell count abnormalities observed in all antibody-related blood cells. These strong alterations may be what impacted some of the reported epigenetic health trait predictions. Of note, according to the developers of those health trait predictors, they should currently only be used at a population level and are not yet supposed to provide reliable predictions at the individual level. Training data from larger-scale cohorts, including diseased individuals, will be required to refine these additional health trait predictors and enable their clinical use^25^. Irrespectively, the clearly abnormal epigenetic measurements and predictions of ID27 would undoubtedly also have led to further clinical tests if detected in a wellness-or preventative healthcare setting, revealing the ongoing but previously undetected pathological process.

Taken together, our findings demonstrate the value and potential of epigenetic predictors and biological age estimators, particularly for risk assessment and early detection. They can reveal unexpected deviations and inform about the probability of having or developing a disease, for example, as shown here with individual ID27. They can also contribute to known clinically relevant outcomes and serve as vital instruments for explaining many biological phenomena. Currently, the extensive and complex preprocessing procedures required to obtain high quality results from omics data are still hindering many of the corresponding predictors from developing their full clinical potential, in particular at the individual level. However, with the field growing and maturing, we argue that BA predictions should be considered as crucial biomarkers that can well-complement routine medical tests and CA.

Indeed, omics data have already begun to enter clinical practice^57^, making omics-based BA predictions feasible. Added to standard laboratory results, they can give a valuable complementary outlook assisting the doctors’ decision-making. Once omics technologies have been fully incorporated into the medical routine, BA predictions will likely become standard measurements regularly discussed between patients and medical professionals. A novel personalised value to identify the “normal” BA for an individual could also be estimated, together with common clinical measures, to provide precise individual interpretations^58,59^. Further longitudinal studies with a more diverse population, including diseased patients, will undoubtedly give broader assurance and validation of the significance of BA predictions at the personal level.

As the field of biological ageing advances, the construction of BA using ageing clocks employing diverse data sources such as epigenomics, proteomics, and metabolomics could be proven effective in uncovering novel biomarkers of biological ageing. The contemporary trend of profiling cohorts through multi-omics technologies is paving the way for a more comprehensive understanding of the molecular underpinnings of ageing. Future endeavours to weave multi-omics into ageing clocks are poised to not only broaden our grasp of the molecular signatures that characterise ageing but also enhance the predictive powers of these models. Furthermore, the successful incorporation of physiological and tissue function parameters into ageing clock models could open new avenues of exploration. The continued expansion of this integrative approach is anticipated to provide more discernible and actionable insights, solidifying the role of ageing clocks as indispensable tools in the evolving landscape of personalised medicine and ageing research.

## Data Availability

All data produced in the present study are available upon reasonable request to the authors.

## Contributors

Conceptualisation, M.P., G.E., and S.E.; Formal analysis, M.P. and S.E.; Visualisation, M.P. and S.E.; Writing – Original draft, M.P., G.E. and S.E.; Writing – Review & Editing, G.E. and S.E.; Supervision; G.E., S.E., O.T., and S.B.

## Acknowledgments

-

## Data sharing

The IAM Frontier longitudinal dataset are available from the corresponding authors on a reasonable request.

## Supplementary Material

A supplementary document is available online.

