## Supplementary Document for "Making Biological Ageing Clocks Personal"

*^*^Corresponding author*

*^†^Equal Contributions*

### 1. DNA methylation data preprocessing

We performed extensive quality assessment of the DNA methylation data using Minfi’s qcPlot and QCReport functions^1^, plotCtrl of the ENmix package^2^ and the QC metrics of Heiss & Just^3^. One out of the 96 samples did not pass the quality control checks and was removed from the dataset. The failed sample was a technical replicate. We also applied a sex check and performed SNP analyses to identify potential sample mixups.^3^ No issues were detected in these analyses.

We filtered probes with a bead count of less than three in at least 5% of samples and probes with a detection p-value > 0.01 in at least one sample. Non-cg probes were also excluded. Probes mapping to sex chromosomes or SNPs were not removed as several epigenetic predictors make use of a subset of these probes. However, they were excluded for data visualization purposes during quality control and preprocessing and specifically accounted for where required, for example during batch effect correction (see below). The probe filtering reduced the number of used probes from 865,859 to 836,477.

We assessed the global distributions of the data using density plots (Supplementary Figure 1), the visualization of PCAs of methylation M-values and MDS plots of methylation Beta-values as well as unsupervised hierarchical clusterings using Euclidean distances and different linkage methods. The raw filtered dataset showed a light batch effect corresponding to the three timepoints of our study but were of good quality otherwise (see Supplementary Figure 1)**.**

All samples from one particular participant of the study were identified as strong global outliers. These were samples from two timepoints and one technical replicate (n=3). The strongly altered DNA methylation profiles did not seem to be caused by technical issues but biological differences and were consistent over time. Therefore, these samples were not removed from the analyses. To avoid biasing the rest of the data because of the three strong outliers during preprocessing steps that borrow information from all samples, we performed our preprocessing pipeline (see below) twice: First, we removed all samples from the corresponding individual for preprocessing. Second, we repeated the same preprocessing procedure now also including the biological outlier samples. We then added the results of the outlier samples preprocessed in the second run back into the main dataset that was preprocessed without the outliers.

Technical replicates from all three timepoints (n=2 per timepoint) and also across timepoints (n=10) were used to assess performance, along with the data visualization procedures as described above. Density plots, PCA, MDS and hierarchical clusterings were repeated after every preprocessing step.

For background correction and dye-bias normalization we applied NOOB (normal-exponential convolution using out-of-band probes).^4^ This method estimates the background mean intensity using out-of-band control probes which provide signals in the opposite fluorescent channel. NOOB effectively adjusts for differences in background distribution and average intensities between samples run on different arrays.^4^ Subsequently, we applied BMIQ (Beta-Mixture Quantile normalization)^5^ to adjust for probe type bias and perform intra-array normalization.

To assess batch effects, we performed singular value decomposition (SVD) of the DNA methylation M-values and Beta-values, excluding probes mapping to the sex chromosomes. The analysis revealed significant batch affects corresponding to array and slide. We employed the empirical Bayesian framework ComBat of the SVA package^6^ to correct for the identified batch affects, using age as phenotype of interest and including sex as covariate as we needed to retain probes mapping to sex chromosomes in our dataset for downstream analyses using epigenetic predictors.

Data visualization as described above and unsupervised hierarchical clustering (see Supplementary Figure 2) confirmed that our preprocessing procedure successfully removed batch effects and samples of the same person always clustered together. Technical replicates also clustered very closely together. Biological subclusters of the same age and sex were visible, while no subclusters of any technical variables were present anymore after normalization and batch effect correction.

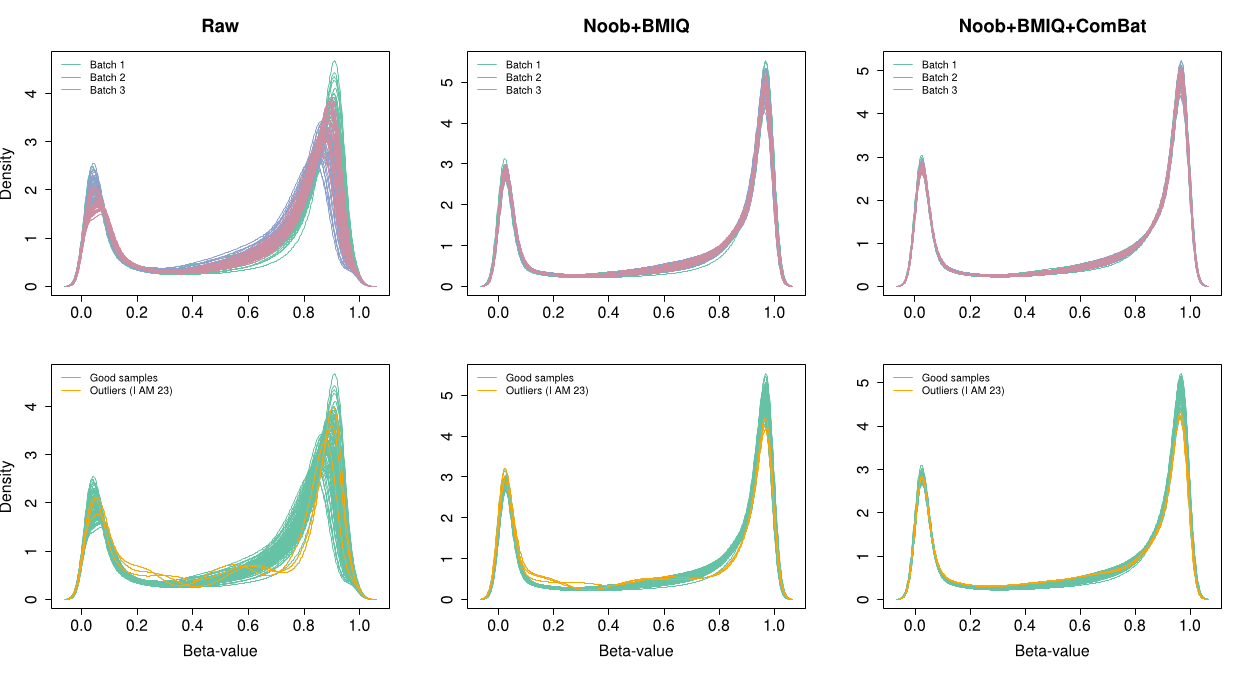

**Supplementary Figure 1**. Density plots of the global distributions before and after normalization and batch-effect correction.

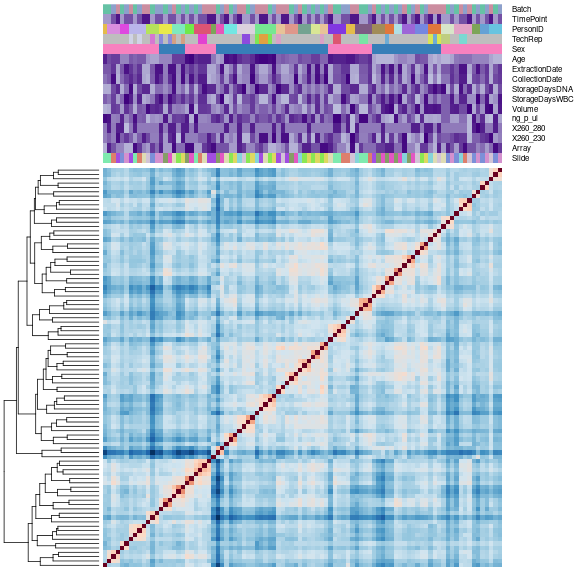

**Supplementary Figure 2**. Unsupervised hierarchical clustering after preprocessing.

### 2. Disease risk scores, proteomics profiles, physical conditions and blood count predictions

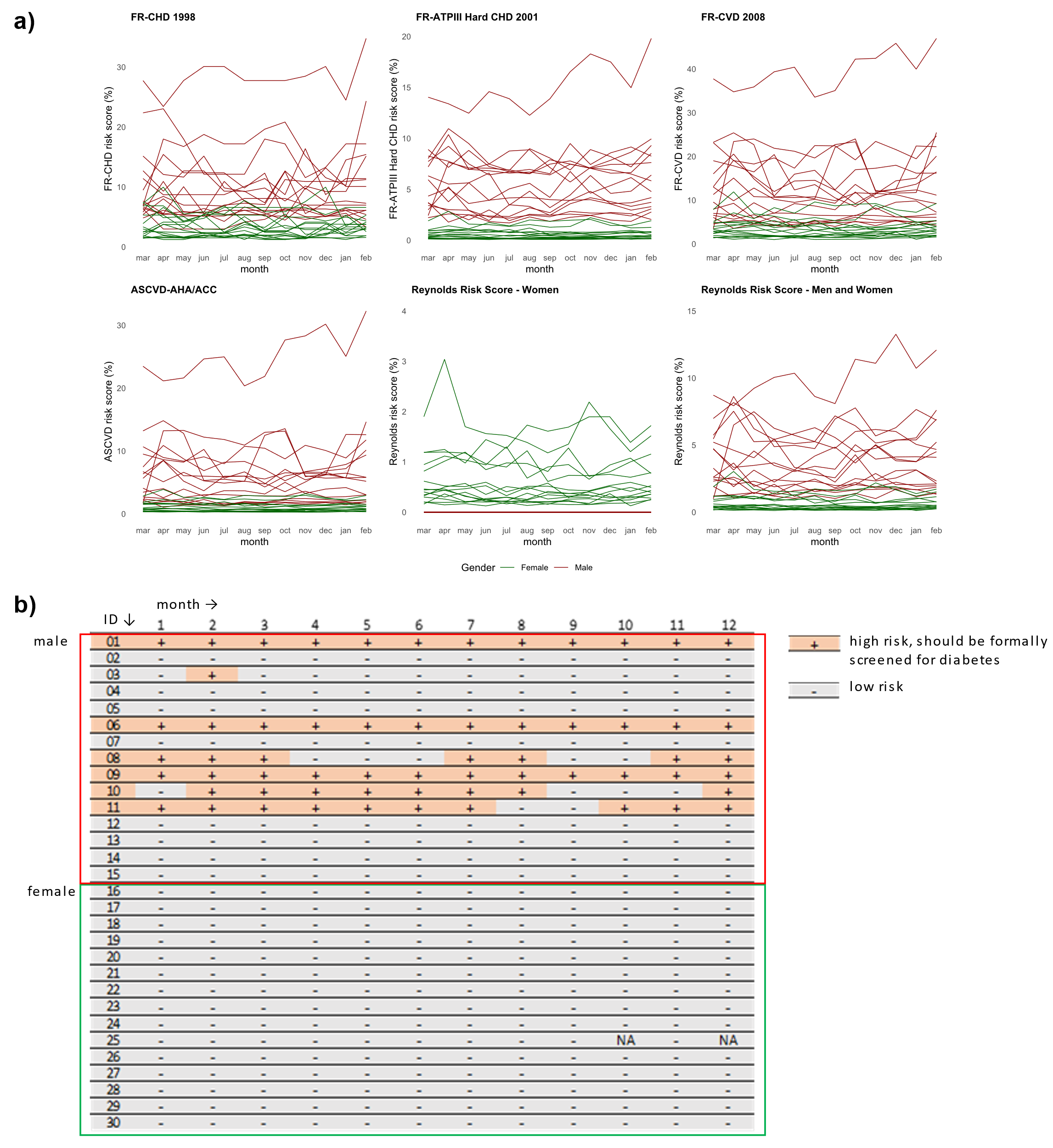

**Supplementary Figure 3**. In every time point during the complete 13-month study period, we computed **a**) 10-year cardiovascular disease (CVD) risk scores^7–12^, and **b**) Diabetes risk scores (calculated based on the American Diabetes Association (ADA) risk calculator^13^.

**a)**
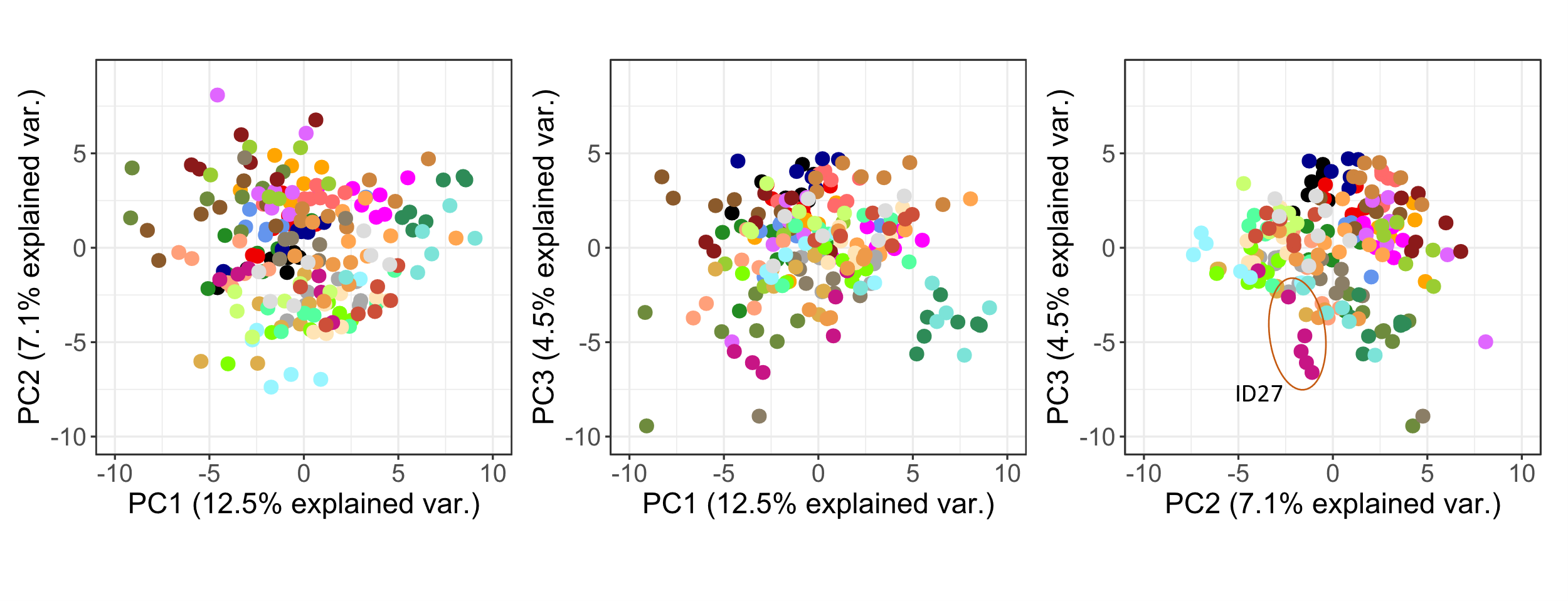

**b)**

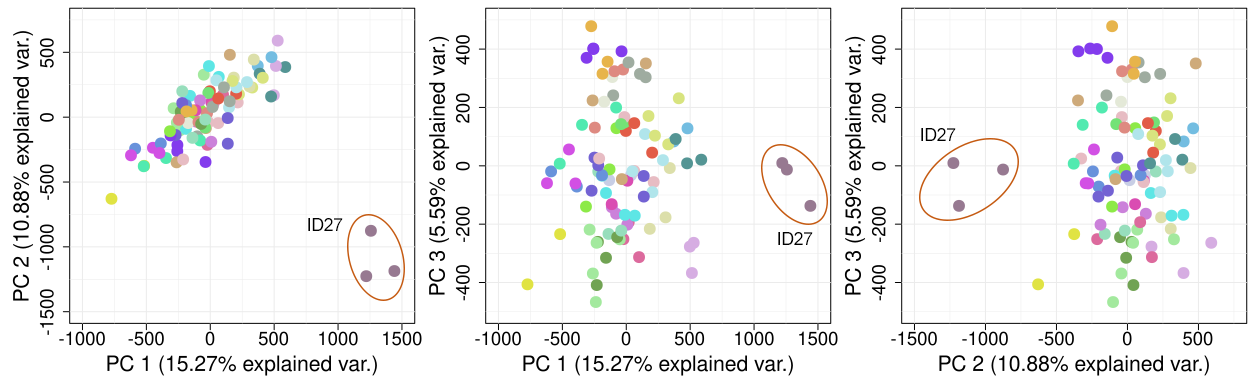

**Supplementary Figure 4**. **a)** Principal component analysis of the proteomics dataset. The explained variabilities were calculated using 1067 protein variables in seven time points. ID27 is found outside the main cluster based on the third principal component. b) Principal component analysis of the preprocessed, normalized and batch effect-corrected DNA methylation dataset containing 836,477 CpGs of 30 individuals across three time points as well as technical replicates (n=95 samples). All samples of ID27 are located outside the main cluster in principal components one and two.

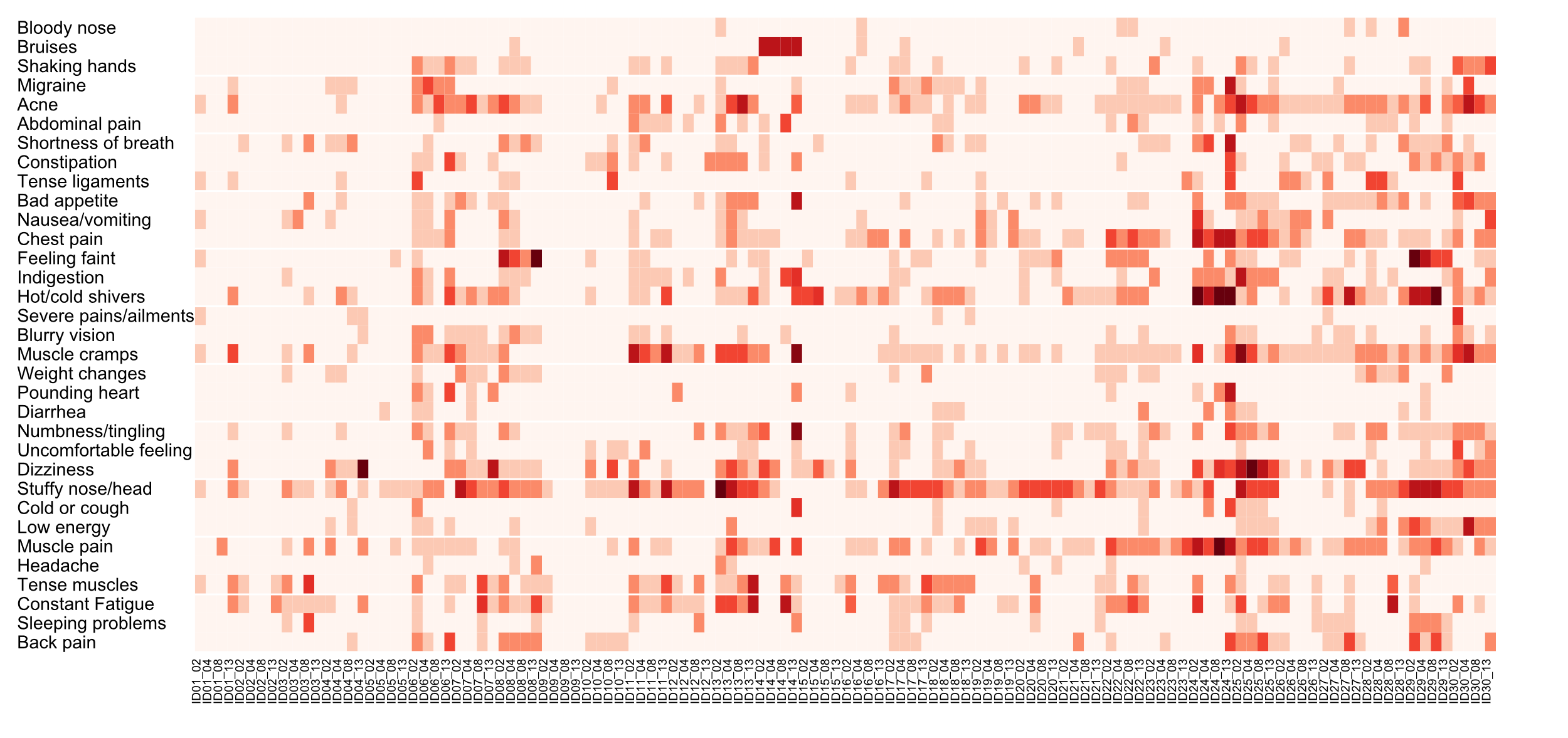

**Supplementary Figure 5**. Heatmap of the physical condition based on weekly questionnaire answers collected during the study. Darker colors indicate that the individuals experienced the condition more often. The numbers after the identifiers indicate the month that matched with the DNA methylation time points; month 2, 4, 8, and 13.

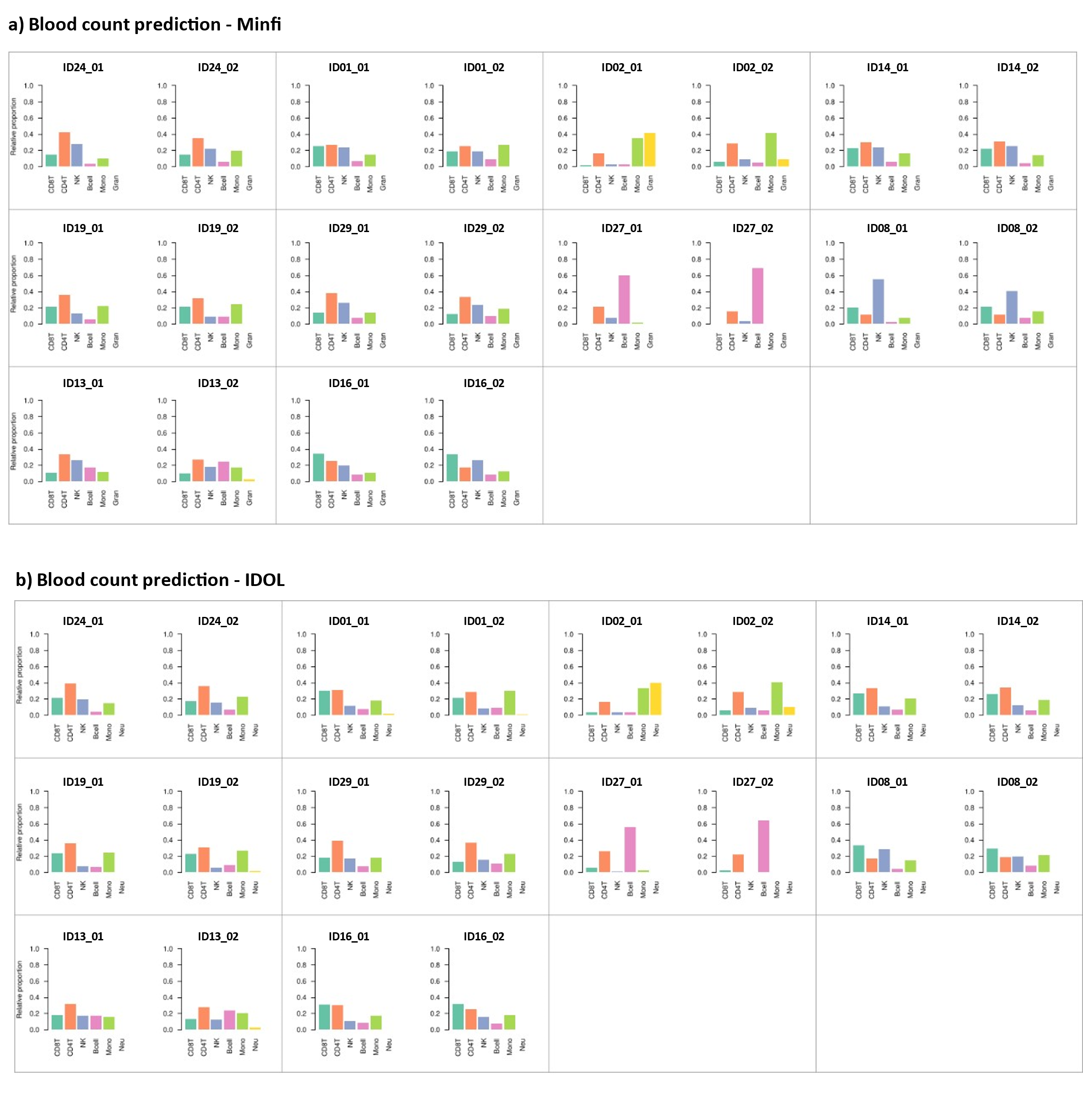

**Supplementary Figure 6.** Blood count predictions estimated by **a)** Minfi and **b)** IDOL in several individuals.
